# Recognizing Activities of Daily Living using Multi-sensor Smart Glasses

**DOI:** 10.1101/2023.04.14.23288556

**Authors:** Simon Stankoski, Borjan Sazdov, John Broulidakis, Ivana Kiprijanovska, Bojan Sofronievski, Sophia Cox, Martin Gjoreski, James Archer, Charles Nduka, Hristijan Gjoreski

## Abstract

Continuous and automatic monitoring of an individual’s physical activity using wearable devices provides valuable insights into their daily habits and patterns. This information can be used to promote healthier lifestyles, prevent chronic diseases, and improve overall well-being. Smart glasses are an emerging technology that can be worn comfortably and continuously. Their wearable nature and hands-free operation make them well suited for long-term monitoring of physical activity and other real-world applications. To this end, we investigated the ability of the multi-sensor OCOsense™ smart glasses to recognize everyday activities. We evaluated three end-to-end deep learning architectures that showed promising results when working with IMU (accelerometer, gyroscope, and magnetometer) data in the past. The data used in the experiments was collected from 18 participants who performed pre-defined activities while wearing the glasses. The best architecture achieved an F1 score of 0.81, demonstrating its ability to effectively recognize activities, with the most problematic categories being standing vs. sitting.

## I. Introduction

Smart sensing with wearable devices has enabled numerous applications in ubiquitous computing. One of the tasks that has emerged as an important pre-requisite for other applications is Human Activity Recognition (HAR). HAR is a broad field of study that identifies a person’s movements or actions based on sensor data. It covers a broad range of activities, from typical indoor activities, such as walking, talking, standing, and sitting, to more focused activities, such as those performed in a kitchen or factory floor [1]. HAR can also be used for tracking transportation modes [2] and stress levels [3], or as a part of disease severity detection methods for various diseases such as Parkinson’s disease and depression monitoring [4]. In healthcare, HAR is used in identifying symptoms of movement disorders or neurological diseases, aiding in diagnosis or symptom management [5].

Historically, collecting sensor data for HAR was a challenging and expensive process, which required custom hardware. This made it difficult to collect and analyze data on a large scale and limited the potential applications of the technology. However, with the widespread availability of smartphones and other wearable devices for fitness and health monitoring, this scenario has changed significantly.

With the recent development of technology, sensor data can be easily obtained, and therefore a larger amount of HAR-based studies can be made.

Body movements are among the main measurable components of behavior and play a significant role in the study of HAR. The use of motion sensor devices, such as accelerometers and gyroscopes, has made it possible for scientists to measure human behavior outside of the laboratory environment. These devices supply researchers with objective and quantitative data based on the three spatial dimensions, making it possible to analyze human movements in detail.

Over the years, a number of wearable devices and sensors have been proposed for automatic detection of everyday activities. Early studies in this field explored the use of sensors placed on different parts of the body, so they can enhance detection accuracy. Over time, the selection of sensors was refined, taking into account two key factors, the capability of the sensors to precisely detect daily activities, and the practicality that encompasses user comfort and acceptability. As a result, the most commonly used devices for HAR are smartphones, wristbands, and smartwatches. However, the recent advancements in technology have enabled the development of compact, lightweight, and stylish smart glasses. Therefore, the researchers in this field started exploring the possibility to detect everyday activities using data coming from sensors embedded in smart glasses [6][7]. This device is suitable for HAR because it can provide an immersive and hands-free experience, enabling the user to perform physical activity without any distractions.

In this study, we investigated the ability of the multi-sensor OCOsense™ smart glasses to recognize everyday activities. For this purpose, we evaluated three end-to-end deep learning architectures that showed promising results when working with sensor data in the past. Additionally, we explored the inter-subject variability for the recorded activities. Finally, we explored how we can include temporal information and detect transitions between activities.

## II. Related Work

The HAR domain has been thoroughly explored in the past using body-worn sensors. Most of the multi-sensor HAR approaches involve machine learning (ML) algorithms (e.g., Random Forest (RF), Support-vector Machines (SVM), and k-Nearest Neighbors (KNN)) to build models from features extracted from each modality of the body-worn sensor. More recently, researchers started exploring novel approaches based on deep learning (DL) for HAR [8][9].

In the field of HAR, sensors are usually placed on the wrists, ankles, hips, waist, or torso of the user. Approaches using head-mounted devices are rather scarce. Loh et al. [10] used a head-worn accelerometer, barometer, and GPS sensors with an SVM for fitness activity classification. Additionally, Zhang et al. [6] and Farooq et al. [11] proposed the use of head-mounted sensors to detect eating and chewing events. Head-AR [12] is a method based on weighted ensemble learning used for HAR from sensors mounted on a VR device.

Despite the limited work in the field, the use of smart glasses for human activity recognition is a promising approach with a unique set of challenges. To this end, Faye et al. [13] published a dataset that is collected with commercially available glasses, smartwatch, and smartphone. The smart glasses provide data from an embedded IMU. This dataset has, however, some noticeable drawbacks. First, only one user participated in the experiment. Moreover, there is no well-defined set of activities or well-defined protocol, which makes it difficult to evaluate or extend.

Another use of Electrooculography (EOG) J!NS MEME glasses have been demonstrated by Ishimaru et al. [14]. The study provides a signal level assessment of MEME glasses and shows the ability to distinguish 4 activities (typing, reading, eating, talking) with an accuracy of 70% for 6 second windows and up to 100% for a 1-minute majority decision.

Meyer et. al [15][16] conducted two studies that propose context-aware human activity recognition (HAR) system for smart glasses by combining eye movement features from laser feedback interferometry (LFI) sensors and head movement features from an IMU. The method presented in his studies was DL based approach using convolutional neural networks (CNNs).

More recently, UCA-EHAR was introduced, and it consists of 20 subjects performing 8 different activities, while wearing Ellcie Healthy (EH) smart glasses equipped with a gyroscope, accelerometer, and barometer, was proposed by Novac et al. [7]. RNN was used for classification of different activities and analysis of the power consumption during live inference on the smart glasses’ microcontroller was performed.

Another effort to develop a system for HAR using smart glasses (Google Glass Explorer Edition XE 22) has been made by Wang et al. [17]. The authors compare the classification performance of a Support Vector Machine (SVM) with data collected either from a smartphone or smart glasses for four activities (biking, jogging, movie watching, and video gaming).

## III. Data

A total of 18 young adult research volunteers (9 males and 9 females) aged between 18 – 37 (23 ± 5) participated in the data collection process, which included individuals engaging in activities of daily living (ADL) in a realistic home environment. There were no strict criteria that needed to be met for individuals to participate in the study. Instead, all individuals who were able to comfortably wear the one-size-fits-all glasses were considered eligible to participate. This approach ensured that the sample size was not limited by factors such as age, gender, or physical ability. Each participant was equipped with emteq’s OCOsense™ smart glasses (Figure *1*), which are fitted with: (i) a 9-axis inertial measurement (IMU) sensor (accelerometer, gyroscope, and magnetometer), (ii) pressure sensor; and (iii) optomyography sensors that measure skin movement. For this study, we used only the data provided by the IMU and pressure sensor. During the data collection procedure, the sensors were continuously sampled at a fixed rate of 50Hz.

**Figure 1.**
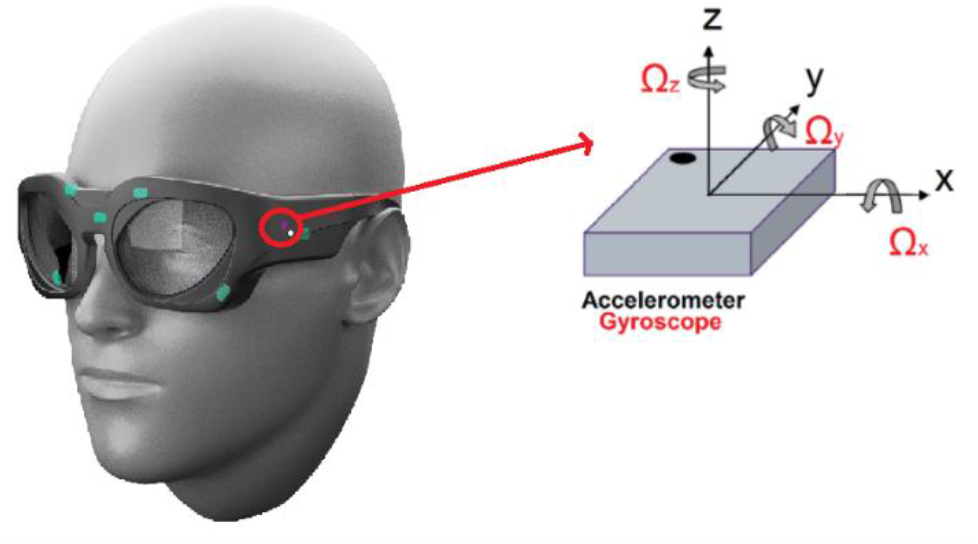
OCOsense™ smart glasses. The green colored rectangles represent the OCO™ sensors. The 9-axis IMU is represented by the purple rectangle.

The data from the smart glasses was streamed in real-time via Bluetooth to an application running on an iPad. The researcher conducting the data collection also performed real-time labeling. To facilitate this, the app featured a screen with buttons representing pre-determined activities. The researcher selected the appropriate button based on the activity being performed by the participant. Additionally, the dataset consists of synchronized video data from five external cameras placed around the apartment, which ensures labeling the activities as accurately as possible.

Prior to the start of the data collection procedure, each participant was instructed on how to wear the glasses properly, and we ensured the fit and comfort. The data collection procedure involved three predefined scenarios that covered the activities of interest.

Each participant was instructed about the scenarios, however, there was no limitation on how to perform a specific activity. The predefined scenarios resulted in three categories of activities:

- Posture activities – included activities related to the posture of the participant, such as sitting, standing, walking, lying and transitions between sitting and standing.
- Personal hygiene activities – included activities that are part of an individual’s daily hygiene routine, namely, washing hands and brushing teeth.
- Dietary intake activities – included activities that are related to nutrient intake, such as eating and drinking.

Each participant was recorded for 2 hours, resulting in a dataset with 36 hours of labelled data. For the purposes of this study, we analyzed the following activities: sitting, standing, lying, walking, eating, drinking and hygiene, which includes washing hands and brushing teeth. The distribution of the data is shown in Figure 2.

**Figure 2.**
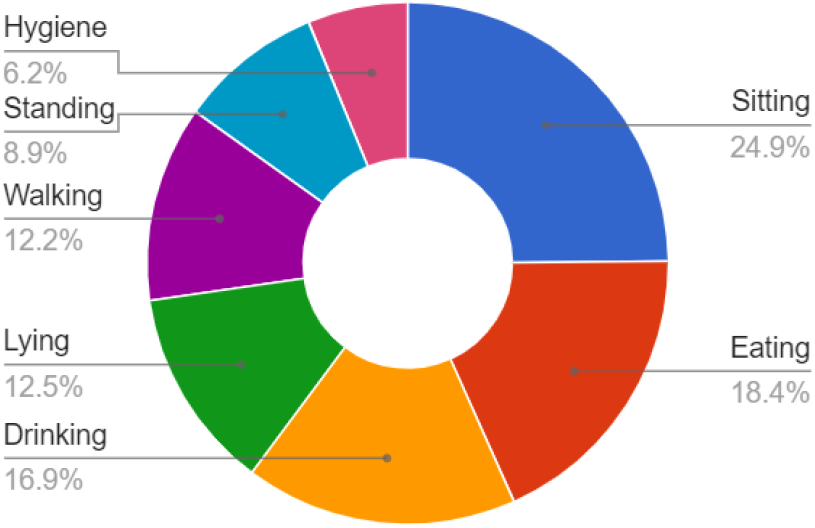
Distribution of the seven activities.

## IV. Method

Before passing the data as an input to the selected DL architectures we filtered the signals using a fifth-order median filter. Once the data was filtered, we generated magnitude for the accelerometer and gyroscope sensors. The next step in the process was to select the optimal window size for windowing the data. We tested various window sizes, and experimentally we determined that window size of 4 seconds contains enough temporal information from where the models can learn the patterns between the different activities. As a result, the signals were segmented using a window size of 4 seconds with a 2 second slide between consecutive windows.

Once the signals were prepared, we used them as an input to three end-to-end DL architectures. The chosen DL architectures are based on well-known architectures used in the past for various problems, such as computer vision and time-series related topics. In this study, the selected architectures were adjusted to work with one dimensional signals.

- Spectro-Temporal Residual Neural Network (STResNet). For each sensor utilizes channel-specific residual blocks and spectrograms to extract temporal and frequency information. This architecture already proved to be successful in previous studies for various applications, such as HAR from smartphone sensors [18], for heart failure detection from heartbeat sounds [19], for monitoring driver distractions from physiological and visual signals [20].
- Inception Network – combines multiple blocks of channel-specific inception modules, which should increase the performance of vanilla CNNs. This performance of this architecture was evaluated in a previous study aimed at detection of eating habits based on smartwatch data [21].
- Feature Level Fusion Network – combines channel-specific CNNs and bidirectional LSTM networks to extract features from the raw sensor data and further aggregates the extracted features by using feature level fusion. This network was successfully implemented in detecting gait abnormalities using data from wrist-worn inertial sensor [22].

For all three architectures the cross-entropy loss was used as the objective function for training. Furthermore, all the models were trained using an Adam optimizer with a dynamic learning rate. The initial learning rate was set to 1e-3 and decreased by a factor of 0.1 every fifty epochs. To avoid overfitting to the training data, we used early stopping callback for all architectures. The callback was set to monitor the F1-macro score of the validation set. If the F1-macro score was not improved over ten epochs the training was stopped. The main reason for monitoring the F1-macro score is because it provides unweighted mean of the F1-scores calculated for each class separately. This way can have a general sense of how well the trained models perform over all classes. For all models NVIDIA Titan Xp GPU was used to accelerate the training process.

## V. Experimental setup and results

To estimate the performance of the previously described DL architectures, we used a 6-fold cross validation (CV) approach. The created folds were participant-based, which means that the data from one participant only appeared in a single fold. The training procedure is repeated 6 times, each time using a different fold as the testing set. By using subject-based k-fold cross-validation, the inter-subject variability of the data is taken into account and helps to reduce the risk of overfitting. To assess the performance of the trained models, we used the F1-macro score and accuracy. The reported results in this study are obtained using predictions for all folds, across all tasks and they are shown as a mean value with the standard deviation between folds.

### A. Performance Evaluation

Table 1 shows the performance of each architecture described in Section IV, including F1-macro score, model size, and the number of parameters. Based on the obtained results, we can see that all architectures show promising results.

**Table 1:**
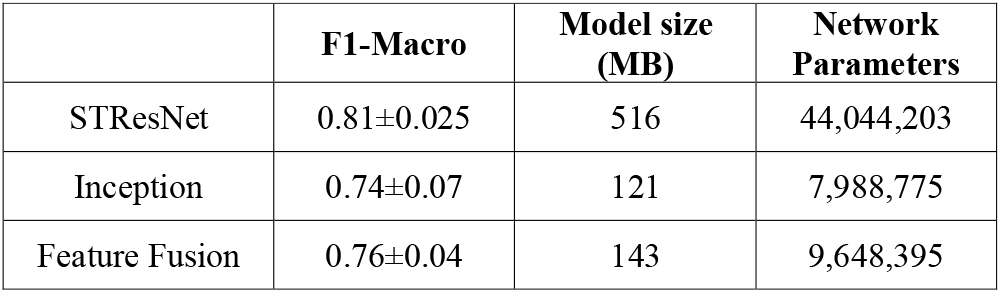
Performance evaluation of each Dl architecture

|  | F1-Macro | Model size (MB) | Network Parameters |
| --- | --- | --- | --- |
| STResNet | 0.81±0.025 | 516 | 44,044,203 |
| Inception | 0.74±0.07 | 121 | 7,988,775 |
| Feature Fusion | 0.76±0.04 | 143 | 9,648,395 |

As shown, STResNet performed the best with an F1-macro score of 0.81. Additionally, the standard deviation of the results obtained with the STResNet architecture is quite small, which suggests that this DL approach can learn general characteristics and perform well on data from unseen participants. Also, obtaining stable results over all participants proves our hypothesis that IMU data from head-mounted device is less prone to movement noise, hence a model based on such data can perform well on new participants. The other two architectures show similar results, however, it can be seen that the Inception architecture is a bit unstable based on the standard deviation between the folds. Additionally, if we compare the F1-macro score of the Inception and Feature Fusion architectures to the STResNet architecture and take into account the size of the models, we can say that the difference in performance is acceptable. To gain better understanding of each architecture performance the confusion matrices are shown in Figure *3*. This analysis provides us with an understanding of the model’s strengths and weaknesses, which can be useful for optimization.

**Figure 3.**
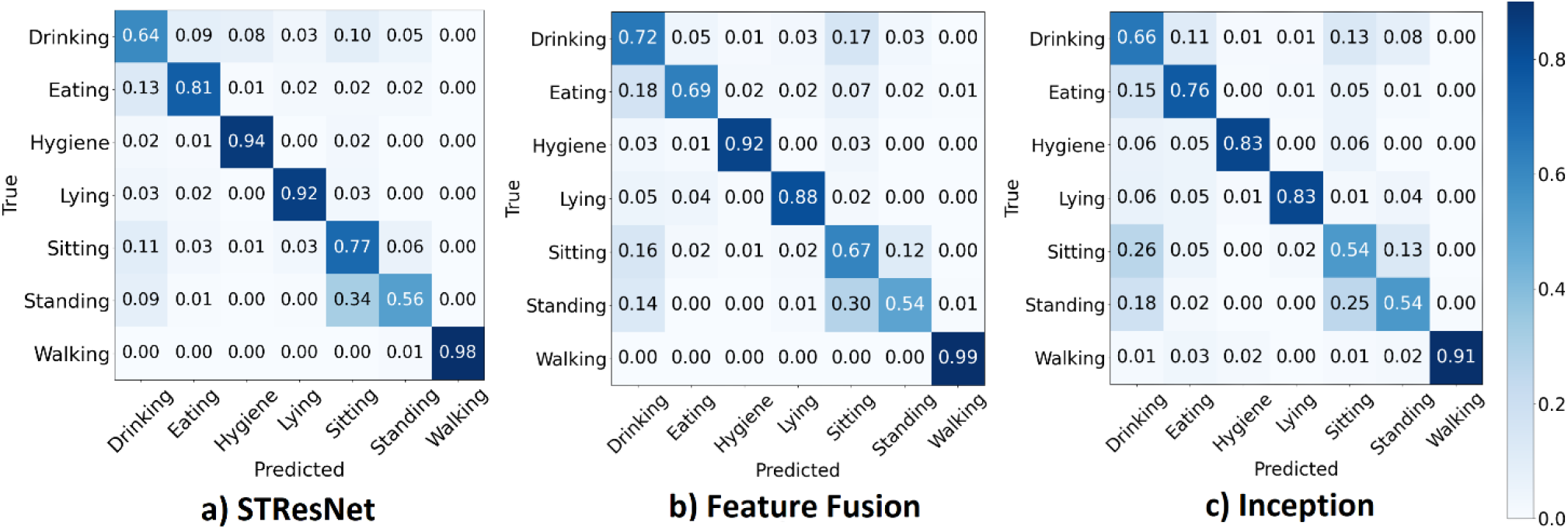
Confusion matrices for each DL HAR model.

For the STResNet architecture, it is evident that the model is able to accurately detect activities such as hygiene, lying, and walking. However, the model faced some challenges when differentiating between standing and sitting. Additionally, in some cases, the model misclassifies eating and drinking activities. The drinking activity sometimes is predicted as sitting and less often as standing.

The Feature Fusion and Inception architectures also exhibit similar limitations as STResNet. Specifically, the Feature Fusion network had a higher misclassification rate for the eating activity. Meanwhile, the Inception architecture had the lowest F1-Macro score for drinking due to a higher rate of misclassifying drinking as sitting. In comparison with STResNet, the misclassification of drinking and eating activities with sitting and standing is noticeably higher.

One of the key issues for all three architectures is the differentiation between standing and sitting. This was in a way expected, given that these two activities resemble each other when looking through the prism of an IMU in smart glasses, i.e., the orientation of the IMU is similar during the two activities. Mixing drinking and eating activities with sitting and standing also arises from this issue. During the drinking activity, the subjects were in a seated position. Additionally, the eating activity was also primarily performed while sitting. This is the cause for misclassifying these activities. Nonetheless, we can see that the STResNet architecture in some situations managed to differentiate between standing and sitting. The reasoning why we expect the models to learn the difference is analyzed in more details in the subsection Exploring Standing vs. Sitting.

### B. Sampling Rate Analysis

The OCOsense™ smart glasses provide sensor data sampled at a fixed rate of 50 Hz. Based on studies done in this field [18], it was established that this sampling frequency is sufficient for most activity recognition tasks. However, high sampling frequency and processing large amount of data can significantly reduce the device’s battery life. To try to mitigate these issues we decided to further evaluate the best performing architecture (STResNet) with down sampled sensor data to 25Hz and 10 Hz.

The results for this experiment are shown in Table 2. It can be clearly seen that the best performance was achieved with 50 Hz data. However, if we compare the results with those achieved when the data was down sampled to 25 Hz, it can be noticed that the performance was decreased minimally. Given the results in similar fields, it’s not surprising that 25 Hz is sufficient for recognizing human motion. Additionally, the training time for the model was reduced by almost one third. The reduced training time of the model with 25 Hz data was also an important consideration, as it allows multiple experiments to be conducted and do analysis faster.

**Table 2:**
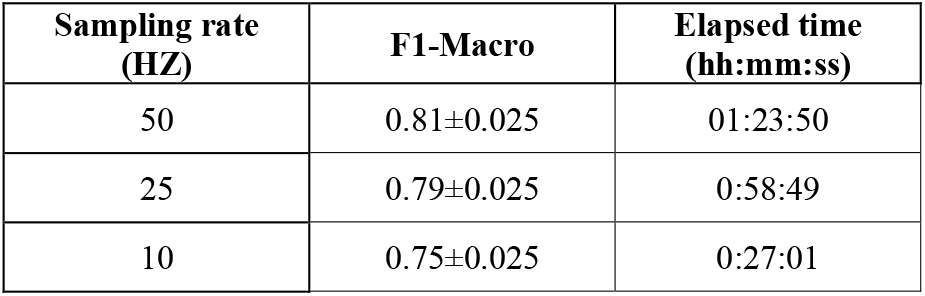
Sampling rates analysis - results

The experiment where the data was down sampled to 10Hz resulted in more significant loss of 0.06 F1-macro score compared to the results where 50 Hz data was used. Nonetheless, these results still show that the model is capable of learning how to differentiate the activities in most of the cases. Similarly as before, by further reducing the sampling frequency to 10 Hz, the training time of the models was reduced to only one third of the original training time when the data was sampled with 50 Hz.

### C. Sensor-specific Experiments

To understand the usefulness of each sensor modality, we investigated the performance of the STResNet architecture by using each modality individually as well as their combinations. The list of sensor modalities tested in this experiment includes data from accelerometer (A), gyroscope (G), magnetometer (M), and pressure (P) sensor. As a result, we had to repeat the training procedure 15 times. Based on the results from the previous section we decided to reduce the training time and worked with 25 Hz data.

The obtained results are shown in Figure 4. The comparison of the results using data from a single sensor modality is colored with blue bars and shows that the accelerometer and the gyroscope are the most informative. Additionally, we can see that when using only data from the pressure sensor we cannot differentiate between activities.

**Figure 4.**
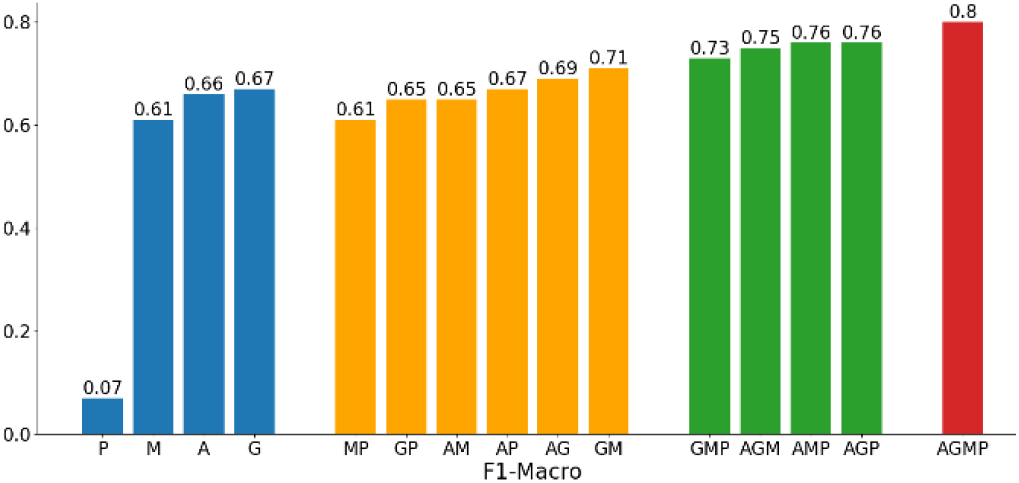
Sensor-specific experiments. A-Accelerometer, G-Gyroscope, M-Magnetometer, P-Pressure sensor.

Furthermore, based on the combinations of data from two sensor modalities, which are colored with yellow, we can see that in certain situations we get improvement over models based on a single sensor modality. The best performing model is based on a combination of data from gyroscope and magnetometer. Next, we have the combination of accelerometer and gyroscope, which also outperforms all single modality results. The remaining combinations did not introduce any improvement over the best performing single sensor modality.

The results where combinations of three sensor modalities are combined are shown with green bars on the plot. Here we can see that in all cases the models based on three sensor modalities outperform all previously discussed combinations. It is interesting to notice that by combining accelerometer, gyroscope, and pressure data the results were improved by 0.07 compared to only accelerometer and gyroscope data combination.

Finally, the red colored bar shown in Figure 4 represents the results when data from all four sensor modalities is combined. Here, it can be clearly seen that model based on all sensor modalities outperforms all previously discussed combinations. Also, we can see that our idea to include pressure data in the model results in improvement, compared to only working with accelerometer, gyroscope, and magnetometer.

### D. Exploring Standing vs Sitting

The confusion matrix presented in Figure *3* showed that the STResNet model wrongly predicted classes standing and sitting. To further investigate the separability of these activities we included transitions from sitting to standing and vice versa and analyzed the raw signal acquired from the pressure sensor.

The result from the analysis is visualized in Figure 5. The different activities are color-coded. The figure shows that the pressure of the subject changes during the four different activities. One interesting observation is that during the transitions “sitting down” and “standing up” the pressure increases and decreases accordingly. Being able to recognize these transitions accurately using data from the pressure sensor, may be useful in distinguishing between standing and sitting activities. Thus, a possible solution for the standing vs. sitting problem would be to have a temporal model that tracks the transitions (“sitting down”, “standing up”). Once we know the last transition of the user, we can more easily infer whether the user is sitting or standing. For example, if the last transition was “sitting down,” then “sitting” is a more probable next activity than “standing.”

**Figure 5.**
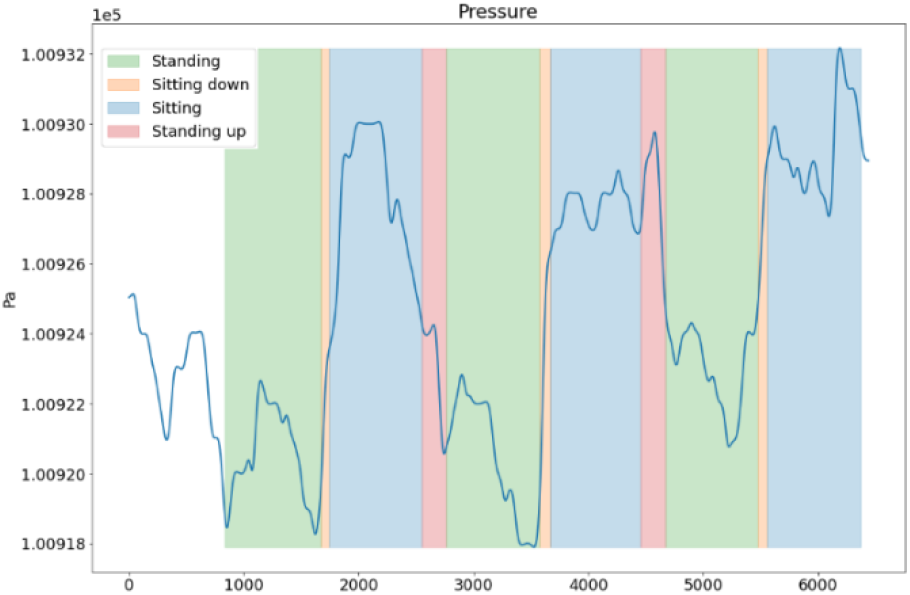
Pressure sensor signal visualization during sitting, standing sitting-to-standing and standing-to-sitting transition data.

## VI. Conclusion

In this study, we analyzed data from the novel OCOsense™ multi-sensor smart glasses and their ability to recognize everyday activities in combination with deep learning models. For this purpose, we collected a HAR dataset that includes common activities of daily living.

We explored three end-to-end deep learning models for HAR, including STResNet, Inception, Feature Fusion models. The best performing model, STResNet, achieved an overall f1-macro score of 0.81 and an accuracy of 0.80. Notably, the model achieved these results without the need for extensive feature engineering in an end-to-end manner, highlighting the power of deep learning in activity recognition.

Additionally, we conducted an experiment where we tested different sampling frequencies. The performance decreased slightly when the data was down-sampled to 25 Hz. A bit larger reduction was obtained when testing with 10 Hz, however, the results were still acceptable.

The analysis where we evaluated the contribution of individual sensor modalities and their combinations showed that the gyroscope is the most informative when working with a single modality. Additionally, this experiment showed that combination of all sensor modalities resulted in the best performance.

Although the presented results in this study are promising, there are still open challenges that we plan to improve in the near future. For instance, the challenge of accurately distinguishing between sitting and standing activities remained present in all experiments. The reason is that the orientation of the head (i.e., the glasses and the IMU) is the same during these two activities. However, we showed that by explicitly analyzing transitions using pressure data we might be able easily distinguish these classes. Additionally, future work for this study can include expanding the dataset to include a more diverse population and a wider range of activities. Furthermore, testing the model in real-world scenarios and evaluating its performance in various environments can provide valuable insights into its practical applicability.

Another important aspect is to investigate the generalization capability of the proposed model on other datasets.

## Data Availability

The data is available on demand.

https://emteq.net/

## ACKNOWLEDGEMENT

This work was supported by NIHR i4i FAST (reference no. 33180) under the project “Context-Aware Multimodal Pervasive Sensing (CAMPS) of behaviour in daily life”, and by Innovate UK under the project “Mobile Observation of Depression (MOOD) platform for digital phenotyping” (Grant number 105207). Hristijan Gjoreski’s work was partially funded by the WideHealth project (EU’s Horizon 2020 research and innovation programme, grant agreement No. 952279).

